# Cerebrovascular disease drives Alzheimer plasma biomarker concentrations in adults with Down syndrome

**DOI:** 10.1101/2023.11.28.23298693

**Authors:** Natalie C. Edwards, Patrick J. Lao, Mohamad J. Alshikho, Olivia M. Ericsson, Batool Rizvi, Melissa E. Petersen, Sid O’Bryant, Lisi Flores-Aguilar, Sabrina Simoes, Mark Mapstone, Dana L. Tudorascu, Shorena Janelidze, Oskar Hansson, Benjamin L. Handen, Bradley T. Christian, Joseph H. Lee, Florence Lai, H Diana Rosas, Shahid Zaman, Ira T. Lott, Michael A. Yassa, José Gutierrez, Donna M. Wilcock, Elizabeth Head, Adam M. Brickman

**Affiliations:** Taub Institute for Research on Alzheimer’s Disease and the Aging Brain, Columbia University, New York City, NY, USA; Department of Neurology, Vagelos College of Physicians and Surgeons, Columbia University, New York City, NY, USA; Department of Neuroscience, Columbia University, New York City, NY, USA; Department of Neurobiology & Behavior, University of California, Irvine, CA, USA; University of North Texas Health Science Center, Fort Worth, TX, USA; Department of Pathology and Laboratory Medicine, University of California Irvine School of Medicine, University of California, Irvine, CA, USA; Department of Neurology, University of California, Irvine, CA, USA; Department of Psychiatry, University of Pittsburgh, Pittsburgh, PA, USA; Clinical Memory Research Unit, Department of Clinical Sciences Malmö, Lund University, Lund, Sweden; Memory Clinic, Skåne University Hospital, Malmö, Sweden; Waisman Center, University of Wisconsin-Madison, Madison, WI, USA; Department of Neurology, Harvard Medical School, Massachusetts General Hospital, Boston, MA, USA; Department of Radiology, Center for Neuroimaging of Aging and neurodegenerative Diseases, Athinoula A. Martinos Center for Biomedical Imaging, Charlestown, MA, USA; Department of Psychiatry, University of Cambridge, Cambridge, UK; Department of Pediatrics and Neurology, School of Medicine, University of California, Irvine, CA, USA; Center for the Neurobiology of Learning and Memory, University of California, Irvine, CA, USA; Stark Neurosciences Research Institute, Indiana University School of Medicine, Indianapolis, IN, USA; Department of Neurology, Indiana University School of Medicine, Indianapolis, IN, USA; Anatomy, Cell Biology and Physiology, Indiana University School of Medicine, Indianapolis, IN, USA

## Abstract

**Importance:** By age 40 years over 90% of adults with Down syndrome (DS) have Alzheimer’s disease (AD) pathology and most progress to dementia. Despite having few systemic vascular risk factors, individuals with DS have elevated cerebrovascular disease (CVD) markers that track with the clinical progression of AD, suggesting a role for CVD that is hypothesized to be mediated by inflammatory factors.

**Objective:** To examine the pathways through which small vessel CVD contributes to AD-related pathophysiology and neurodegeneration in adults with DS.

**Design:** Cross sectional analysis of neuroimaging, plasma, and clinical data.

**Setting:** Participants were enrolled in Alzheimer’s Biomarker Consortium – Down Syndrome (ABC-DS), a multisite study of AD in adults with DS.

**Participants:** One hundred eighty-five participants (mean [SD] age=45.2 [9.3] years) with available MRI and plasma biomarker data were included. White matter hyperintensity (WMH) volumes were derived from T2-weighted FLAIR MRI scans and plasma biomarker concentrations of amyloid beta (Aβ42/Aβ40), phosphorylated tau (p-tau217), astrocytosis (glial fibrillary acidic protein, GFAP), and neurodegeneration (neurofilament light chain, NfL) were measured with ultrasensitive immunoassays.

**Main Outcomes and Measures:** We examined the bivariate relationships of WMH, Aβ42/Aβ40, p-tau217, and GFAP with age-residualized NfL across AD diagnostic groups. A series of mediation and path analyses examined causal pathways linking WMH and AD pathophysiology to promote neurodegeneration in the total sample and groups stratified by clinical diagnosis.

**Results:** There was a direct and indirect bidirectional effect through GFAP of WMH on p-tau217 concentration, which was associated with NfL concentration in the entire sample. Among cognitively stable participants, WMH was directly and indirectly, through GFAP, associated with p-tau217 concentration, and in those with MCI, there was a direct effect of WMH on p-tau217 and NfL concentrations. There were no associations of WMH with biomarker concentrations among those diagnosed with dementia.

**Conclusions and Relevance:** The findings suggest that among individuals with DS, CVD promotes neurodegeneration by increasing astrocytosis and tau pathophysiology in the presymptomatic phases of AD. This work joins an emerging literature that implicates CVD and its interface with neuroinflammation as a core pathological feature of AD in adults with DS.

**Key Points:** *Question:* Do white matter hyperintensities, a magnetic resonance imaging marker of small vessel cerebrovascular disease, predict plasma Alzheimer’s biomarker concentrations of amyloid, tau, and neuroinflammatory pathophysiology and downstream neurodegeneration in adults with Down syndrome?

*Findings:* Increases in white matter hyperintensity volume precede and promote inflammation- and tau-related pathophysiology, directly and indirectly, leading to downstream neurodegeneration.

*Meaning:* Small vessel cerebrovascular disease may contribute to the pathophysiological progression of Alzheimer’s disease in adults with Down syndrome. These findings support the hypothesis that cerebrovascular disease is a core feature of Alzheimer’s disease in adults with Down syndrome.

## Introduction

Virtually all individuals with Down syndrome (DS) develop Alzheimer’s disease (AD) pathology, including abnormal amyloid-beta (Aβ) plaques and tau neurofibrillary tangles, by the age of 40 years^1,2^ and most develop dementia by the age of 60.^3^ Down syndrome is considered a genetic form of AD^4^ and pathogenesis is attributable to the triplication of chromosome 21, which contains the amyloid precursor protein (*APP*) coding gene.^5^ Models of AD progression in both late onset and genetic forms emphasize the role of Aβ in initiating tau pathology and subsequent neurodegeneration, sometimes referred to as the ‘ATN framework.’^6^ While there is general support for this pathophysiological cascade,^6,7^ there is increasing evidence that additional pathways may promote AD pathogenesis and progression.^8^

Cerebrovascular disease contributes to risk and course of clinical AD and increases the likelihood of developing dementia.^9,10^ Neuroimaging biomarkers for small vessel CVD, including white matter hyperintensities (WMH), are associated with neurodegeneration, indexed by AD-related patterns of cortical atrophy and fluid biomarker concentrations.^11–13^ Despite consistent observations of its occurrence and contributions to clinical outcomes in people with AD, CVD is generally considered a common comorbidity with AD that is not a hallmark characteristic of the disease.^6^

We turned to populations at genetic risk to determine the extent to which CVD represents a ‘core feature’ of AD. Among community-dwelling older adults, those carrying the *APOE* ε4 allele, the strongest genetic risk factor for late onset AD, there are greater degrees of cerebrovascular disease than in non-ε4 carriers.^14^ Despite their younger age and relatively low vascular risk factor profiles, individuals with autosomal dominant, fully penetrant mutations for AD, have increased WMH volumes up to 20 years prior to expected symptom onset compared with individuals without genetic mutations for AD but who are at similar risk.^13^ Such changes account for more variance in cognition than do other AD biomarkers.^15^ Similarly, individuals with DS generally have lower degrees of vascular risk compared with neurotypical adults and seem to be protected against developing hypertension,^16–18^ yet have neuroimaging evidence of CVD that increases with clinical progression of AD.^19^

Evidence from late onset and genetic forms of AD suggests that cerebrovascular pathology is indeed a prominent feature of AD that cannot be attributable solely to exposure to vascular risk factors, but whether CVD promotes primary AD pathophysiological progression remains unclear. Reports of associations between CVD and AD biomarkers are mixed, with some showing codependency^20^ and others not.^21,22^ In a preclinical model of WMH, we found that white matter damage induced by transient hypoperfusion promotes tau hyperphosphorylation, but it is unclear what factors mediate this effect.^12^ Emerging work suggests the critical role of neuroinflammation, mainly manifesting as a change in microglia morphology,^23–26^ astrocytosis,^27–29^ and inflammatory mediators,^30^ in AD pathogenesis and course, with emerging evidence of intimate crosstalk between inflammatory processes and the brain’s vasculature.^31^ In adults with DS, MRI markers of CVD are associated with proteomic patterns reflective of inflammation earlier in the disease and with patterns reflective of neurodegeneration later in the disease.^32^ Glial fibrillary acidic protein (GFAP) is a cytoskeletal protein found in astrocytes, released during astrogliosis, and can be measured reliably in cerebrospinal and blood compartments.^33^ GFAP concentration is elevated in people with and at-risk for AD^34–36^ and appears to mediate the relationship between Aβ and tau pathology.^37,38^ In adults with DS, plasma GFAP concentration discriminates between individuals who are asymptomatic and those diagnosed with AD.^39^

Further, GFAP levels are strongly correlated with indicators of Aβ and tau pathology, neurodegeneration, and clinical progression of AD in adults with DS.^39–41^

In the current study, we examined the association between WMH, as a marker of small vessel cerebrovascular disease, and AD plasma biomarker concentrations, including Aβ40/Aβ42, phosphorylated tau 217 (p-tau217), and GFAP, with neurofilament light chain (NfL) across disease stages in adults with DS. Because (i) astrocytosis is prominent around blood vessels in AD,^42^ (ii) induced cerebral hypoperfusion, a characteristic of DS,^43^ increases the number of GFAP positive astrocytes,^44^ and (iii) astrocytosis is an early disease feature of AD,^45^ we used a series of mediation and path analyses to test our hypothesis that CVD gives rise to tau pathology and ultimately neurodegeneration via astrocytosis across different AD disease stages in adults with DS.

## Methods

### Participants and participant diagnosis

Participants came from the Alzheimer Biomarkers Consortium – Down Syndrome (ABC-DS), a multisite, observational study designed to examine biomarker, clinical, and genetic correlates of and contributors to AD among adults with DS.^46^ The sample included individuals from the Neurodegeneration in Aging Down Syndrome (NiAD; U01 AG051406) and Biomarkers of Alzheimer’s Disease in Adults with Down Syndrome (ADDS; U01 AG051412), both of which are now contained within ABC-DS. For the current study, participants with available MRI data and derived plasma biomarkers of interest were selected for analysis. One hundred thirty-eight participants characterized as cognitively stable, 24 patients with mild cognitive impairment (MCI), 16 patients with AD dementia (DS-AD), and 8 with diagnoses that were “unable to be determined” were included. Diagnoses were based on a consensus conference that reviewed available neuropsychological and clinical data, as described previously in detail.^46^ In short, clinical experts in the assessment and diagnosis of AD in DS performed a standardized clinical evaluation of each participant, which considered functional abilities and health history. Participants were then assigned one of four AD-related diagnoses (i.e., cognitively stable, MCI, DS-AD, unable to be determined). This study was conducted in accordance with the institutional review boards of participating institutions and written informed consent was obtained from each participant or their legal guardian or legally authorized representative.

### Magnetic resonance imaging

MRI scans were acquired at ADDS and NiAD participating sites. NiAD sites acquired 2D T2-weighted fluid-attenuated inversion recovery (FLAIR) scan (repetition time [TR]/ echo time [TE]/ inversion time [TI] = 5,000/386/1,800 milliseconds, voxel size = 0.4 × 0.4 × 0.9mm^3^) and ADDS sites acquired 3D T2-weighted FLAIR scan (TR/TE/ TI = 4,800/119/1,473 milliseconds, voxel size = 0.9 × 0.9 × 0.5mm^3^).

White matter hyperintensity volume was quantitated with in-house software. Briefly, FLAIR images were reconstructed to a uniform matrix of 256×256×256 with a voxel size of 1 mm^3^. The images were reoriented to standard anatomical space (MNI152), skull stripped, and bias field corrected.^47,48^ The images were processed through a customized module designed to extract percentile thresholds from the intensity histogram of each image automatically.^49,50^ Next, a white matter segment was created using the convolutional neural networks tool.^51^ Two specific percentile thresholds were computed: one for the transition between dark and bright voxels intensity, and another for the transition between bright and brightest voxels intensity. These thresholds initialized a Gaussian mixture model (GMM) and expectation-maximization algorithm^52^ within the white matter segment of the FLAIR images, using two components to represent hyperintense and non-hyperintense voxels.

Following the computation of percentile thresholds, we calculated the inter-percentile range (IPR) between these values and introduced a relaxed threshold of 10 to account for variations in FLAIR image quality. This adjustment was made by applying a multiplicative factor to the IPR.

Finally, probability distribution maps were generated to represent the segmented WMH within the FLAIR images. The Roberts edge detection function^53^ was applied to the probability distribution maps, ensuring the removal of any non-white matter voxels from the brain’s contour. The labeled voxels were added together and multiplied by voxel dimensions to calculate total WMH volume in cm^3^. **Figure 1** displays the voxel-wise frequencies of WMH across all participants.

**Figure 1.**
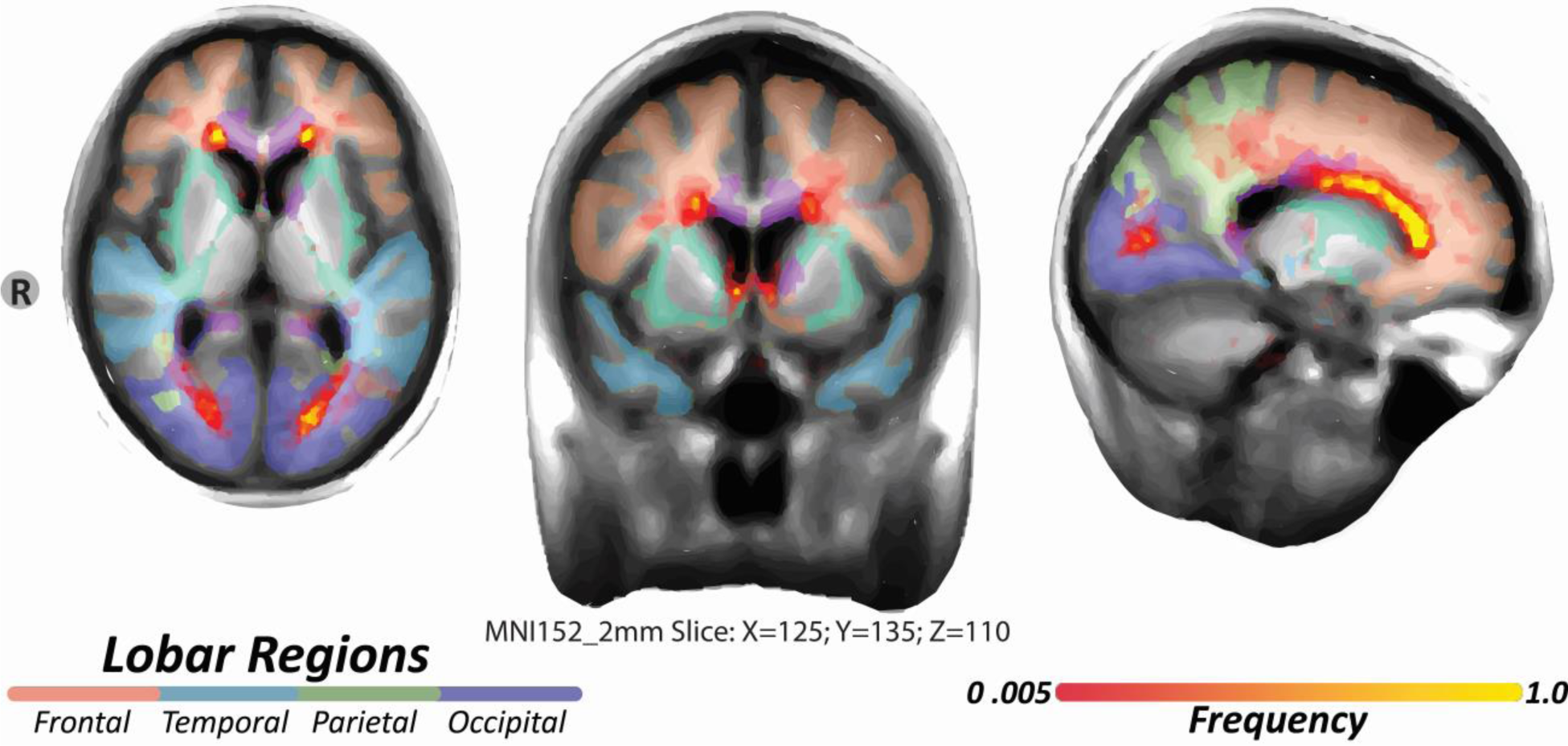
Frequency map of white matter hyperintensities in adults with Down syndrome. A voxel-wise frequency map of WMH was created by summing voxels labeled across all 185 individual 3D and dividing by 185. Each voxel’s value represents the proportion of times it was labeled as a WMH across the 185 masks from low frequency (red) to high frequency (yellow).

### Plasma samples and analysis

Plasma Aβ42, Aβ40, p-tau217, NfL, and GFAP concentrations were derived for each participant from plasma samples as previously described.^40^ Plasma samples were shipped to the University of North Texas where Aβ42, Aβ40, and NfL concentrations were quantified with single molecule array (Simoa) assays (Quanterix). We calculated the ratio of Aβ42 to Aβ40 as the biomarker for amyloid pathology.^54^ Plasma samples from the same group of participants were shipped to Lund University for quantification of p-tau217 and GFAP concentrations. The p-tau217 concentration was assayed according to the published protocols using immunoassay on a Mesoscale Discovery platform developed by Lilly Research Laboratories as previously descibed.^41,55^ GFAP concentration was quantified using Simoa assays (Quanterix). We calculated age-residualized values for NfL concentration values, our primary dependent variable, because the pathophysiological progression of AD among individuals with DS is strongly age-dependent.^1,2^ This age-dependency may induce epiphenomenological relationships among AD-related variables when conducting cross-sectional analyses due to their shared association with age. On the other hand, statistical adjustment for age may obscure important associations among factors whose variance is strongly age dependent. Therefore, we chose to operationalize neurodegeneration as age-residualized NfL in the subsequent analyses.

### Statistical Analysis

#### Association of biomarkers with neurodegeneration

We examined the association of WMH and each ADRD biomarker concentration with age-residualized NfL in the entire sample and stratified by diagnosis with bivariate Pearson correlations. Participants with an undetermined diagnosis were not included in any analyses stratified by diagnosis.

#### Mediation analyses

We conducted a series of causal mediation analyses in the entire sample of the observed bivariate associations. We used the ‘mediation’ package in R^56^ to examine whether 1) GFAP mediates the relationship between WMH volume and p-tau217 concentration, 2) whether p-tau217 concentration mediates the relationship between WMH volume and NfL concentration, and 3) whether p-tau217 concentration mediates the relationship between GFAP and NfL concentrations. To probe directionality, we ran models in which the hypothesized predictor and mediator were switched. The average causal mediation effect (ACME), the portion of the direct effect on the outcome that is attributable to the mediator’s effect, and the corresponding p-value were extracted from each mediation model.

#### Path analysis

We tested our *a priori* hypothesis of pathophysiological cascade that is initiated by CVD with a path analysis in the combined sample and in groups stratified by AD-related diagnosis. The path analysis tested the effect of WMH on downstream neurodegeneration (i.e., age-residualized NfL concentrations) via GFAP and p-tau217. The paths were estimated using the ‘lavaan’ package in R,^57^ which established the direct and indirect effects between biomarkers and the model was fit using the sem() function. All analyses were adjusted for research site.

## Results

Sample characteristics across diagnostic groups are reported in **Table 1**. Cognitively stable participants and those with an undetermined diagnostic status were younger than those with MCI and DS-AD and a lower proportion of women were diagnosed with MCI than other groups. There were no differences in reported history of hypertension, hypotension, type 1 or 2 diabetes, or hypercholesterolemia (**Table 1**).

**Table 1.** Sample characteristics by diagnostic group. APOE: apolipoprotein E, MCI: mild cognitive impairment.

|  | **Cognitively Stable** | **MCI** | **Dementia** | **Undetermined** | **Whole Sample** | **Test Statistic** | **p-value** |
| --- | --- | --- | --- | --- | --- | --- | --- |
| ***n*** | 137 | 24 | 16 | 8 | 185 |  |  |
| ***Demographic*** |  |  |  |  |  |  |  |
| *Age, mean (SD) years* | 43 (9.1) | 51 (5.8) | 54.5 (6) | 48.4 (8.8) | 45.2 (9.3) | $F = 27.05$ | <0.001 |
| *Women, n (%)* | 64 (47) | 4 (17) | 9 (56) | 4 (50) | 81 (44) | $\chi^2 = 8.7$ | 0.003 |
| ***Vascular risk factors*** |  |  |  |  |  |  |  |
| *Hypertension, n (%)* | 3 (2) | 0 (0) | 0 (0) | 0 (0) | 3 (2) | $F = 0.9$ | 0.08 |
| *Hypotension, n (%)* | 3 (2) | 1 (4) | 0 (0) | 0 (0) | 4 (2) | $F = 0.8$ | 0.06 |
| *Type 1 diabetes, n (%)* | 0 (0) | 0 (0) | 0 (0) | 0 (0) | 0 (0) | -- | -- |
| *Type 2 diabetes, n (%)* | 2 (2) | 0 (0) | 0 (0) | 0 (0) | 2 (1) | $F = 0.6$ | 0.1 |
| *Hypercholesterolemia, n (%)* | 15 (11) | 1 (4) | 2 (13) | 1 (13) | 19 (1) | $\chi^2 = 1.1$ | 0.3 |

We confirmed strong associations between age and AD biomarker concentrations, apart from Aβ42/40: GFAP: r=0.612 [0.513, 0.695], p<0.0001; NfL: r=0.523 [0.409, 0.62], p<0.0001; p-tau217: r=0.379 [0.248, 0.496], p<0.0001; Aβ42/40: r=0.107 [−0.036, 0.248], p=0.147.

### Associations of plasma biomarkers and WMH with age-residualized NfL concentration across diagnostic groups

**Table 2** displays the associations of plasma biomarkers and WMH volume with age-residualized NfL concentration. WMH volume, GFAP concentration, and p-tau217 concentration were positively associated with age-residualized NfL in the entire sample. In cognitively stable participants, neither WMH volume nor plasma AD biomarker concentrations were associated with age-residualized NfL levels, likely reflecting the limited amount of variance in these factors at this disease stage. Among those with MCI, increased WMH volume was associated with higher age-residualized NfL, while increased GFAP concentration and p-tau217 concentration was associated with higher age-residualized NfL in participants with MCI and AD.

**Table 2.** Associations of plasma biomarkers and WMH with age-residualized NfL concentrations across diagnostic groups.

|  | Whole Sample |  | Cognitively Stable |  | MCI |  | Dementia |  | Undetermined |  |
| --- | --- | --- | --- | --- | --- | --- | --- | --- | --- | --- |
|  | r [CI] | p-value | r [CI] | p-value | r [CI] | p-value | r [CI] | p-value | r [CI] | p-value |
| WMH volume | 0.32<br>[0.18, 0.44] | <0.0001 | 0.14 [-0.03, 0.30] | 0.10 | 0.62<br>[0.23, 0.80] | 0.003 | 0.42<br>[0.18, 0.85] | 0.12 | 0.44 [-0.39, 0.87] | 0.32 |
| Aβ42/40 | 0.14 [-0.01, 0.28] | 0.06 | 0.12 [-0.05, 0.28] | 0.18 | -0.14 [-0.52, 0.28] | 0.51 | 0.31 [-0.22, 0.70] | 0.24 | -0.41 [-0.87, 0.41] | 0.31 |
| p-tau217 | 0.52<br>[0.41, 0.62] | <0.0001 | 0.10 [-0.07, 0.26] | 0.25 | 0.86<br>[0.70, 0.94] | <0.0001 | 0.82<br>[0.55, 0.94] | <0.0001 | 0.21 [-0.58, 0.80] | 0.61 |
| GFAP | 0.31<br>[0.18, 0.44] | <0.0001 | 0.09 [-0.081, 0.25] | 0.31 | 0.47<br>[0.08, 0.74] | 0.02 | 0.62<br>[0.25, 0.87] | 0.005 | 0.19 [-0.59, 0.79] | 0.65 |

*Aβ : β-amyloid, WMH: white matter hyperintensities, p-tau217: phosphorylated tau 217, GFAP: glial fibrillary acidic protein, NfL: neurofilament light chain, MCI: mild cognitive impairment*

### Mediation analyses

Three results emerged from the statistical mediation analyses. First, p-tau217 concentration mediated the relationship between WMH and age-residualized NfL concentration (ACME[CI]=0.44 [0.17, 0.83], p<0.0001). When we reversed the independent variable and mediator variable and re-ran the analyses, WMH did not mediate an association between p-tau217 concentration and NfL (ACME[CI]=0.88 [−0.66, 2.50], p=0.22). Second, GFAP concentration mediated the relationship between WMH and p-tau217 concentration (ACME[CI]=0.0201 [0.01, 0.02], p<0.001); the reverse model revealed a congruent mediation effect of WMH, albeit to a lesser extent, on the relationship between GFAP and p-tau217 (ACME[CI]=0.0003 [0.0001, 0.0002], p<0.001). Third, p-tau217 concentration mediated the relationship between GFAP concentration and age-residualized NfL concentration (ACME[CI]=0.04 [0.01, 0.05], p<0.0001). When reversed, GFAP did not mediate an association between p-tau217 concentration and NfL (ACME[CI]=-0.80 [−2.96, 2.66], p=0.67). A *post-hoc* analysis revealed an interaction between WMH and GFAP on p-tau217 concentration, such that WMH was most strongly associated with p-tau217 in the presence of elevated GFAP while GFAP was most strongly associated with p-tau217 in individuals with high WMH volume (see **Figure 2**).

**Figure 2.**
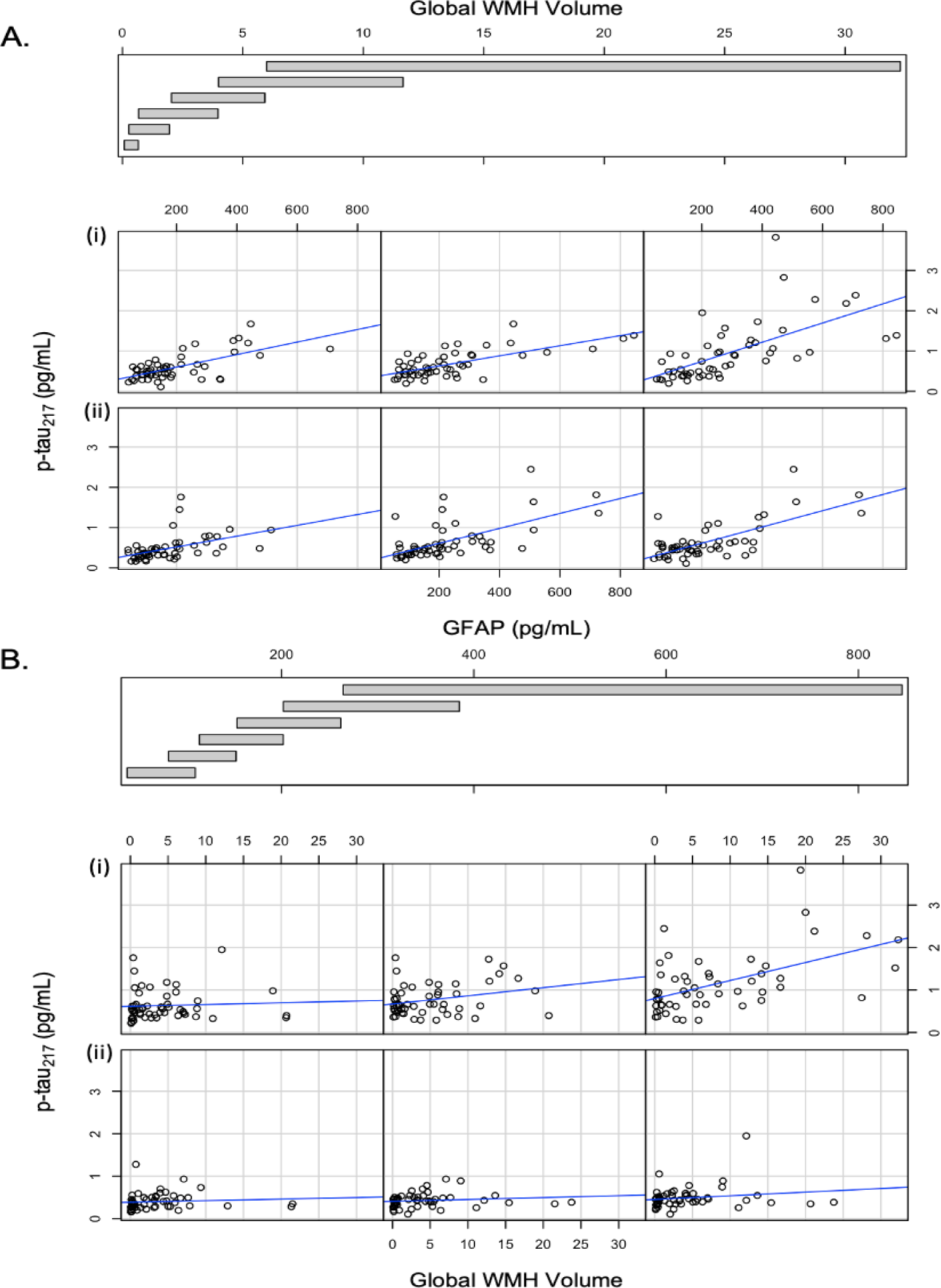
Conditional relationship between WMH and GFAP on p-tau217 concentration. Relationship between GFAP and p-tau217 concentration conditioned by WMH volume (A) and relationship between WMH and p-tau217 concentration conditioned by GFAP (B). The plots show the relationship between GFAP or WMH and p-tau217 for different ranges of WMH and GFAP, respectively. The panels are read from bottom left to top right along each row with the bottom row representing the lowest range of WMH volume and the top row representing the highest range of WMH volume. Rows demonstrating the relationship in individuals with higher distributions are indicated by (ii) while relationships in participants with lower distributions are indicated by rows labeled (i). For example, in Figure 2A., the top right plot shows the relationship between GFAP and p-tau217 in individuals with the largest WMH volume (i) while the bottom left panel shows the relationship between GFAP and p-tau217 in individuals with the smallest WMH volume (ii). WMH: white matter hyperintensities, GFAP: glial fibrillary acidic protein, p-tau217: phosphorylated tau 217

### Path analysis

Informed by the relationships observed in the mediation analyses, we conducted a path analysis to examine statistical causality within our hypothesized pathophysiological cascade in the entire sample and stratified by diagnosis. In the entire sample (**Figure 3A**), the analysis revealed a cascade initiated by WMH, which had a direct and an indirect effect through GFAP on p-tau217 concentration. P-tau217 concentration, in turn, was associated with age-residualized NfL concentration. In this combined sample, increasing p-tau217 concentration was primarily attributable to increasing WMH volume, while increases in NfL were mainly related to increasing p-tau217 concentration. Among cognitively stable participants (**Figure 3B),** there was a direct and indirect effect through GFAP of WMH on p-tau217 concentration, but p-tau217 concentration was not associated with NfL concentration. No AD biomarkers were associated with age-residualized NfL concentrations among cognitively stable participants, likely due to low variance in neurodegeneration at this disease stage. Among those with MCI **(Figure 3C**), increased WMH volume had a direct effect on p-tau217 and NfL concentrations but not GFAP concentration. GFAP concentration had an indirect effect on NfL concentration through p-tau217. Finally, in those diagnosed with dementia **(Figure 3D**), there were no direct or indirect effects of WMH on plasma AD biomarker concentrations. Still, GFAP continued to have a positive indirect effect on NfL through p-tau217 concentration.

**Figure 3.**
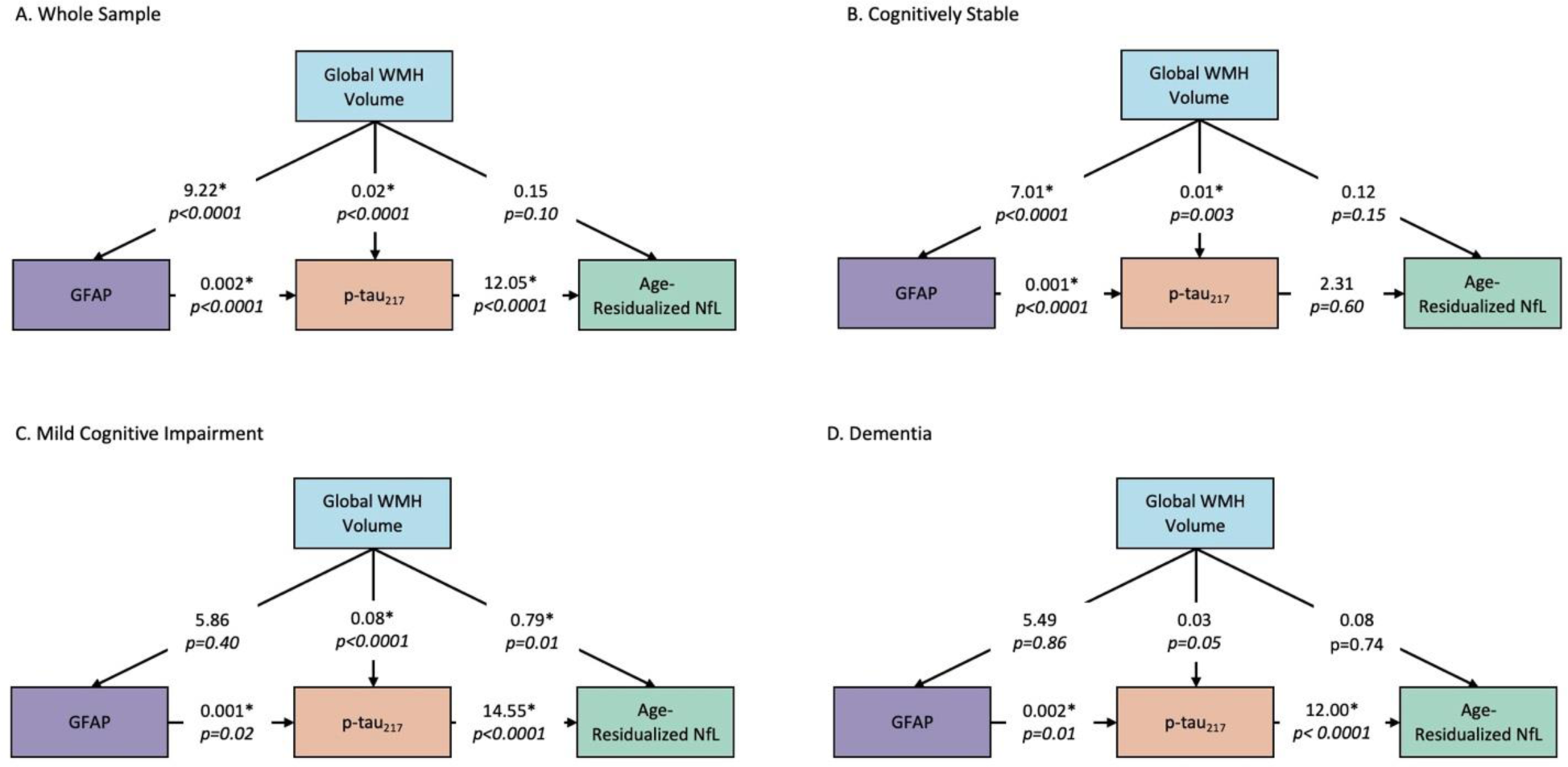
Path models for biomarker progression across diagnostic groups. Statistical modeling calculates relative causal relationships among different pathophysiological contributors. Larger numbers (regression coefficients) signify stronger direct effects. Aβ: β-amyloid, WMH: white matter hyperintensities, p-tau217: phosphorylated tau 217, GFAP: glial fibrillary acidic protein, NfL: neurofilament light chain, MCI: mild cognitive impairment.

## Discussion

Our findings suggest that among adults with DS, CVD promotes AD-related neurodegeneration indirectly through increasing astrocytosis and tau pathophysiology in the preclinical stages of the AD and directly and indirectly in the clinical stages of AD. These results support our hypothesis that WMH may initially promote increases in inflammation and tau pathophysiology, giving rise to downstream neurodegeneration.

Pathogenic models of AD emphasize a precipitating role of Aβ that leads to tau pathology and subsequent neurodegeneration;^6^ however, we found that tau pathology did not appear to exert a direct effect on neurodegeneration until elevated by both cerebrovascular disease and astrocytosis. *Post-hoc* analyses revealed an interaction between GFAP and WMH in promoting AD pathophysiology and downstream neurodegeneration, suggesting synergy between vascular and inflammatory processes in this pathophysiological cascade.

Our findings are consistent with our previous study, which showed more consistent associations between peripheral proteomic markers of inflammation and MRI markers of cerebrovascular disease in presymptomatic phases of AD among adults with DS.^32^ Further, postmortem data revealed a unique inflammatory profile in adults with DS and inflammatory proteins related to astrocytosis were elevated in the early stages of AD.^58^ Adults with DS also show evidence of blood-brain barrier (BBB) disruption at autopsy.^59^ Individuals with DS may be genetically predisposed to BBB disruption, which may promote the cerebrovascular lesions observed on MRI.^60^ Additionally, in mouse models of AD, white matter abnormalities, including those due to hypoperfusion,^12^ promote AD pathology^61,62^ and astrocytosis is an early event in AD in both humans and mouse models.^45^ Therefore, it is possible that BBB dysruption gives rise to cerebrovascular lesions, which has a subsequent impact on astrocytosis, downstream tau accumulation, and neurodegeneration.

White matter hyperintensities are generally considered to reflect “end organ” ischemic damage due to chronic exposure to vascular risk factors.^63^ In the context of AD, however, the etiology of WMH has been widely debated.^63^ Some argue that in AD WMH are attributable primarily to cerebral amyloid angiopathy (CAA).^64^ Others argue that WMH are the *result* of AD-related neurodegeneration, so called Wallerian degeneration.^65,66^ We have argued against some of these pathways on the basis of temporality, experimental evidence, and anatomical distribution.^67^ The results of this study provide further evidence against these possibilities. First, as noted, adults with DS have minimal vascular risk factors; for example, only 2% of participants had a history of hypertension. Nonetheless, WMH are observed across every disease stage. Second, in *post hoc* analyses we did not observe an association between WMH volume and number of cerebral microbleeds (r=-0.043 [−0.226, 0.143], p=0.649), a radiological marker of CAA.^19,68^ This observation is consistent with our finding that cerebral microbleeds only modestly mediate an association between autosomal dominant mutations for AD and increased WMH volume.^69^ Finally, our statistical modeling suggests that WMH precede or are upstream from tau pathophysiology and neurodegeneration markers. Animal stroke models show evidence of increased GFAP positive astrocytes observed around the lesion days after vessel occlusion, specifically in models of small vessel and white matter stroke^70,71^ and white matter hypoperfusion increases tau hyperphosphorylation in mouse models.^12^ Taken together, we speculate that there is an endogenous cerebrovascular component to AD pathogenesis that likely is not associated with amyloid and tau pathology but instead interacts with inflammatory processes to promote tauopathy and subsequent neurodegeneration.

Notably, we did not find any association between plasma Aβ concentration and WMH volume or NfL concentration. This finding was unexpected, given the well-documented overproduction of Aβ in individuals with DS.^1^ However, plasma Aβ concentrations remain steady across the adult lifespan in adults with DS after about age 30 years^72^ and may have plateaued in most participants prior to enrollment. Therefore, the lack of dynamic range in Aβ concentrations may have yielded null results despite the importance of the amyloid pathology. Additionally, plasma amyloid measures may not capture brain-related amyloid pathology with as high fidelity as the other plasma AD biomarkers.^73,74^

## Conclusions

Our study provides evidence of a pathophysiological cascade reflecting a progression of AD pathology in adults with DS initiated by vascular pathology and suggests that cerebrovascular disease and inflammation play a key role and early in AD-related neurodegeneration in adults with DS. As these individuals do not have the same vascular risk factors as the neurotypical population, our study suggests an endogenous vascular component implicated in the progression of AD in DS. The associations between cerebrovascular disease and AD plasma biomarkers in this study indicate that abnormal vascular pathology is a core disease feature and could be a critical treatment target for the DS population.

## Data Availability

Qualified investigators can submit requests for access to data and samples (https://pitt.co1.qualtrics.com/jfe/form/SV_cu0pNCZZlrdSxUN), and all requests will be reviewed by ABC-DS investigators and NIH staff.

## Acknowledgements

The Alzheimer’s Biomarkers Consortium–Down Syndrome (ABC-DS) is funded by the National Institute on Aging and the National Institute for Child Health and Human Development (U01 AG051406, U01 AG051412, U19 AG068054). The work contained in this publication was also supported through the following National Institutes of Health Programs: The Alzheimer’s Disease Research Centers Program (P50 AG008702, P30 AG062421, P50 AG16537, P50 AG005133, P50 AG005681, P30 AG062715, and P30 AG066519), the Eunice Kennedy Shriver Intellectual and Developmental Disabilities Research Centers Program (U54 HD090256, U54 HD087011, and P50 HD105353), the National Center for Advancing Translational Sciences (UL1 TR001873, UL1 TR002373, UL1 TR001414, UL1 TR001857, UL1 TR002345), the National Centralized Repository for Alzheimer Disease and Related Dementias (U24 AG21886), and DS-Connect® (The Down Syndrome Registry) supported by the Eunice Kennedy Shriver National Institute of Child Health and Human Development (NICHD). In Cambridge, UK this research was supported by the NIHR Cambridge Biomedical Research Centre and the Windsor Research Unit, CPFT, Fulbourn Hospital Cambridge, UK.

The authors are grateful to the ABC-DS study participants, their families and care providers, and the ABC-DS research and support staff for their contributions to this study. This manuscript has been reviewed by ABC-DS investigators for scientific content and consistency of data interpretation with previous ABC-DS study publications. The content is solely the responsibility of the authors and does not necessarily represent the official views of the NIH, the CPFT, the NIHR or the UK Department of Health and Social Care

## Author Contributions

*Concept and design:* NCE, AMB

*Analysis, acquisition, and interpretation of data:* NCE, PJL, MJA, MEP, SO, LFA, DLT, OH, BTC, EH, AMB

*Statistical analysis:* NCE, PJL, AMB

*Drafting of the manuscript:* NCE, AMB

*Critical revision of the manuscript:* NCE, PJL, MJA, OME, BR, MEP, SO, LFA, SS, MM, DLT, SJ, OH, BLH, BTC, JHL, FL, HDR, SZ, ITL, MAY, JC, DMW, EH, AMB

^#^**Alzheimer’s Biomarker Consortium-Down Syndrome (ABC-DS) Investigators:** Howard J. Aizenstein, MD PhD; Beau M. Ances, MD PhD; Howard F. Andrews, PhD; Karen Bell, MD; Rasmus M. Birn, PhD; Adam M. Brickman, PhD; Peter Bulova, MD; Amrita Cheema, PhD; Kewei Chen, PhD; Bradley T. Christian, PhD; Isabel Clare, PhD; Ann D. Cohen, PhD; John N. Constantino, MD; Eric W. Doran, MS; Natalie C. Edwards, BS; Anne Fagan, PhD; Eleanor Feingold, PhD; Tatiana M. Foroud, PhD; Benjamin L. Handen, PhD; Jordan Harp, PhD; Sigan L. Hartley, PhD; Elizabeth Head, PhD; Rachel Henson, MS; Christy Hom, PhD; Lawrence Honig, MD; Milos D. Ikonomovic, MD; Sterling C Johnson, PhD; Courtney Jordan, RN; M. Ilyas Kamboh, PhD; David Keator, PhD; William E. Klunk, MD PhD; Julia K. Kofler, MD; William Charles Kreisl, MD; Sharon J. Krinsky-McHale, PhD; Florence Lai, MD; Patrick Lao, PhD; Charles Laymon, PhD; Joseph H. Lee, DrPH; Ira T. Lott, MD; Victoria Lupson, PhD; Mark Mapstone, PhD; Chester A. Mathis, PhD; Davneet Singh Minhas, PhD; Neelesh Nadkarni, MD; Sid O’Bryant, PhD; Melissa Parisi, MD PhD; Deborah Pang, MPH; Melissa Petersen, PhD; Julie C. Price, PhD; Margaret Pulsifer, PhD; Michael S. Rafii, MD PhD, Eric Reiman, MD; Batool Rizvi, MS; Herminia Diana Rosas, MD; Laurie Ryan, PhD; Frederick Schmitt, PhD; Nicole Schupf, PhD; Wayne P. Silverman, PhD; Dana L. Tudorascu, PhD; Rameshwari Tumuluru, MD; Benjamin Tycko, MD PhD; Badri Varadarajan, PhD; Desiree A. White, PhD; Michael A. Yassa, PhD; Shahid Zaman, MD PhD; Fan Zhang, PhD.

